# SARS-CoV-2 seroconversion in response to infection and vaccination: A time series local study in Brazil

**DOI:** 10.1101/2022.03.10.22271805

**Authors:** Luciano F. Huergo, Nigela M. Paula, Ana C.A. Gonçalves, Carlos H.S. Kluge, Paulo H.S.A. Marins, Haxley S.C. Camargo, Thamyres P. Sant’Ana, Lucas R.P. Farias, Juliane D. Aldrighi, Ênio S. Lima, Guiomar T. Jacotenski, Letícia R. Vargas, Gisele Costa, Karin V. Weissheimer, Maria G. Nazário, Kádima N. Teixeira, Marcelo S. Conzentino

## Abstract

The investigation of antibodies raised against different SARS-CoV-2 antigens can help to determine the extent of previous SARS-CoV-2 infections in the population and track the humoral response to vaccination. Therefore, serological surveys can provide key information to better manage the pandemic and/or to implement the most effective vaccination program. Here we describe a time series anti-Nucleocapsid, anti-Spike IgG serological survey analysis in the city of Matinhos, PR, Brazil during the year of 2021. Seroconversion rates to the Nucleocapsid antigen was not influenced by gender or age. Comparison of the serological data with official COVID-19 cases in the city suggest that case sub notification is higher than 47%. Furthermore, by applying serological data, the corrected infection fatality rate was estimated to be lower than 2.4 % in contrast with the official estimative of 3.6 %. The rates of IgG reactive to Spike antigen resembled the curve of the fraction the population that had taken the second vaccine dose. Up to 82% of Spike seroconversion was detected in the end of 2021 confirming the effective of the COVID-19 vaccination program in the city. This SARS-CoV-2 serological study unraveled the SARS-CoV-2 infection rates and the response to vaccination in the city of Matinhos. It is likely that the numbers reported here may be similar in other cities in Brazil.

## Introduction

The COVID-19 pandemic has caused profound impacts in human health throughout the world. The first SARS-CoV-2 case was reported in Brazil by the end of February 2020. In less than two years official numbers indicate that SARS-CoV-2 had infected more than 23 million resulting in > 613.000 deaths in the Brazilian territory https://coronavirus.jhu.edu/map.html. In the state of Paraná, COVID-19 spread rapidly through all cities including Matinhos, the site of this study, where the first case was reported in March 2020. With a population of 35.705 (https://www.ibge.gov.br/cidades-e-estados/pr/matinhos.html) the city of Matinhos had reported, by November 2021, 4.052 COVID-19 cases and 147 deaths https://www.saude.pr.gov.br/Pagina/Coronavirus-COVID-19.

Following the world spread of SARS-CoV-2, international efforts were directed towards the development of an effective vaccine. The results of these intensive global efforts came shortly by the end of 2020 when the first vaccines became available to the public. By January 2022, more than 10 billion doses have been administered worldwide https://ourworldindata.org/covid-vaccinations. In Brazil, the vaccination program started on January 2021 and more than 300 million doses have been administered in one year. By January 2022, 63.000 vaccine doses have been applied to the citizens of Matinhos https://www.saude.pr.gov.br/Pagina/Coronavirus-COVID-19.

The COVID-19 vaccines available in Brazil were CoronaVac – Sinovac/Butantan; AstraZeneca-Oxford - AZD1222 ChAdOx1; Pfizer-BioNTech - BNT162b2 and Janssen - JNJ-78436735 Ad26.COV2.S, all these vaccines have been subjected to extensive clinical trials which confirmed their safety and efficacy (Kim, Marks and Clemens 2021). Despite this, antivaccine groups remain active in Brazil using social media to spread misinformation regarding the safety and efficacy of COVID-19 vaccines.

Serological analysis has been considered as an effective strategy to determine the prevalence of COVID-19 in the population. By tracking long lasting IgG reactive to SARS-CoV-2 antigens it is possible to confirm previous COVID-19 cases which were not notified for different reasons which may include: lack of testing, wrong time of sampling to detect active infection or unnoticed asymptomatic infections (Huang *et al*. 2020a; Petherick 2020). Serological surveys can provide important information to track the evolution of the pandemic, allowing incidence estimates at the population level. Serological analysis can also be used to track the humoral response to vaccination (Huang *et al*. 2020b; Long *et al*. 2020; Petherick 2020). With the estimation of the humoral response raised by the vaccine’s health authorities can determine the extent of the population that has raised antibodies and for how long these antibodies are going to last. Such information is critical to optimize resources and develop an effective vaccination program. In this work we report a year-round SARS-CoV-2 serological analysis in the city of Matinhos, Paraná, Brazil. This study provided insights into the population seroconversion in response to infection and vaccination.

## METODOLOGIA

### Study design and sampling

A local COVID-19 seroprevalence survey was conducted between January to December 2021 in the City of Matinhos, Paraná, Brazil. Participants were enrolled for the study by advertisement at university web site, radio and television. Ethics approval was obtained from the CEP/UFPR (n# 35872520.8.0000.0102). Informed consent was obtained from all participants. A questionary was presented online (the questionary was optional in the beginning of the study but became mandatory to since June 2021). In the questionary participants provided personal information: age, gender and the city of residence. We initially attempted to add a self-declaration for the participant to inform a previous qRT-PCR COVID-19 positive diagnose. However, as different types of COVID-19 tests became available during this study (antigen and serological), most participants were not able to distinguish between the different types of tests. Therefore, a simpler form was used in which the participant could self-declared a previous COVID-19 diagnose and informed the date of symptoms onset. During this study, vaccination against COVID-19 became available so we included a self-declaration questionary for COVID-19 immunization which became mandatory to fill since June 2021. Participants had to indicate if they had been vaccinated, the vaccine manufacture’s popular name (CoronaVac, AstraZeneca, Pfizer or Janssen), the date of the first, second and third doses (if any).

Sampling campaigns were performed at Federal University of Paraná in the city of Matinhos. Approximately 0.1 ml of blood was collected by finger puncture. Samples were centrifuged (5,000 x g for 3 min) and four microliters serum was used to investigate IgG reactive against the SARS-CoV-2 Nucleocapsid or full prefusion Spike using a high throughput magnetic immunofluorescent assay described previously (Conzentino *et al*. 2021b; Huergo *et al*. 2021). The details of antigen purification and bead preparation have been described previously (Alvim *et al*. 2020; Conzentino *et al*. 2021a). The results of each sample were reported and the % of a reference and the method operated with specificity and sensitivity of >99.5% and >95% respectively (Conzentino *et al*. 2021b) .

Confidence intervals were calculated using the online application https://www.surveysystem.com/sscalc.htm. The population of the city of Matinhos was estimated in 35,705 https://www.ibge.gov.br/cidades-e-estados/pr/matinhos.html and prevalence values of 10 or 50 percentage were used to obtain confidence intervals for Nucleocapsid and Spike IgG analysis, respectively. The seroprevalence results were compared with official numbers of reported COVID-19 cases and deaths occurring in the city of Matinhos and with the official numbers of applied vaccines in the State of Paraná. These data were available from the Paraná State Secretary of Health (SESA-PR). https://www.saude.pr.gov.br/Pagina/Coronavirus-COVID-19. The estimated population of the Paraná state was 11,597,484 and the vaccine eligible population (>12 years old) in 2021 was estimated to be 8,883,672 based on the population pyramid predictions by the Brazilian Institute of Geography and Statistics IBGE https://www.ibge.gov.br/apps/populacao/projecao/box_piramideplay.php?ag=41.

## Results

A total of 1384 samples were collected from 982 different individuals. All samples were analyzed for the presence of IgG reactive to SARS-CoV-2 Nucleocapsid protein. Since July 2021 samples were also evaluated for the presence of IgG reacting to SARS-CoV-2 full length Spike protein (736 samples). Altogether 2120 serological analysis were performed. The age of the volunteers had a mean of 44 years (SD 13.5). The age distribution for the participants is presented in Fig. 1A.

**Fig 1.**
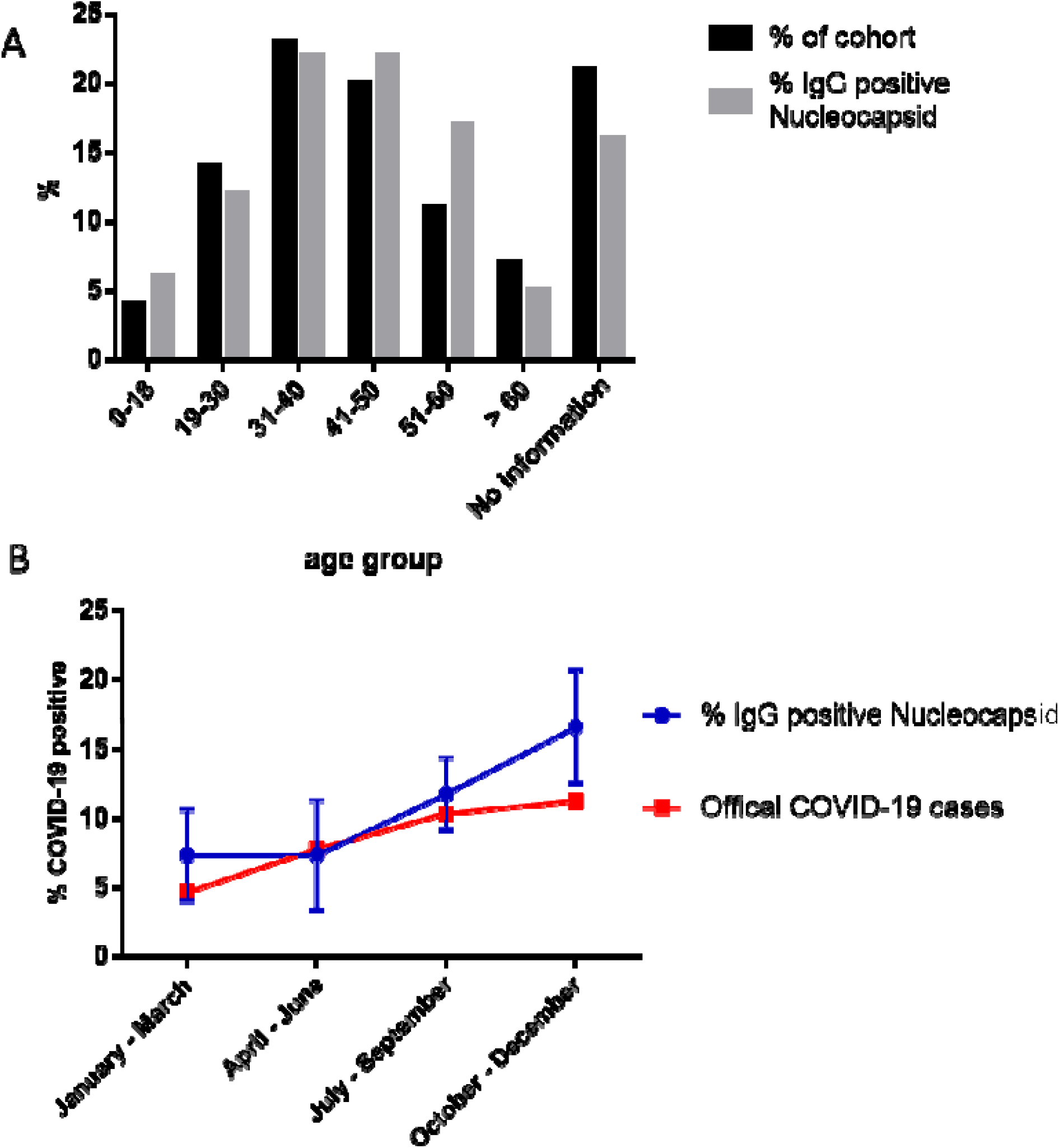
COVID-19 cases detected by IgG reactive to Nucleocapsid antigen. A) The % age distribution of the cohort is shown as black bars. % IgG positive to SARS-CoV-2 nucleocapsid antigen by age group is shown as grey bars. B) Evolution of COVID-19 cases during each trimester of 2021. The % of positive samples for IgG reactive to SARS-CoV-2 nucleocapsid antigen is represented by the blue line. Errors bars indicate the confidence intervals at 95% confidence level. The red line indicates the % of official COVID-19 cases reported by health authorities in the city of Matinhos (accumulated cases in the middle of each trimester is reported as % of total population).

Of the 1384 samples analyzed for IgG reacting to SARS-CoV-2 Nucleocapsid protein 154 were positive 11.1% positive rate (95% CI 9.6 – 12.6). The distribution of the positive cases by gender were similar. Women represented 64% of the cohort and 66% of the positive cases. On the other hand, men represented 36% of the cohort and 34% of the positive cases. The distribution of positive cases by age group also resembled the distribution of the cohort (Fig. 1A).

Vaccination against COVID-19 became available in Brazil by the end of January 2021. Among the vaccines available was CoronaVac representing 24.5% of the vaccines distributed in Brazil. Given that CoronaVac is based on inactivate SARS-CoV-2 it could elicit humoral response against the SARS-CoV-2 nucleocapsid antigen in such way that it would not be possible to distinguish if a nucleocapsid positive IgG test would be caused by previous SARS-CoV-2 infection or due to CoronaVac vaccination.

To evaluate the impact of CoronaVac vaccination in our dataset we evaluated SARS-CoV-2 nucleocapsid seroconversion rate after the exclusion of Coronavac vaccinated participants (excluded samples N =180). Quite surprisingly, the nucleocapsid IgG positive rate was not significantly affected (11.1% vs 10.9% after exclusion). Indeed, we noticed that most subjects vaccinated with CoronaVac tested negative for IgG reactive to Nucleocapsid protein. Of the 63 samples whose donors declared not to be previously diagnosed with COVID-19 and to have taken the second Coronavac dose more than 10 days, only 12 % were positive (mean time between second dose and sampling - 73 days, SD 69). For all subsequent analysis considering nucleocapsid IgG data the Coronavac vaccinated subjects were excluded. Hence, all remaining nucleocapsid IgG positive cases would be caused only by previous SARS-CoV-2 infections.

The IgG reacting to Nucleocapsid seroconversion rate accordingly to the time frame of the study is depicted on Fig 1B. The samples were categorized accordingly to the trimester of sampling during 2021. In the first trimester of the study (January to March) 7.4% of samples were positive for nucleocapsid IgG. The same number was detected in the second trimester (April to June); 11.8 in the third trimester (July to September) finally reaching 16.7% in the fourth trimester (October to December). Despite the limitation of sample size in each trimester, which incur in broad confidence intervals, the data suggests that SARS-CoV-2 infections has continued to increase during the time frame of the study (Fig. 1B). The rate of nucleocapsid seroconversion followed the trend of official COVID-19 cases in the city of Matinhos (Fig. 1B). The reported official accumulated COVID-19 cases in the middle of each trimester are within the confidence intervals of the number of cases detected by nucleocapsid seroconversion, expected in the fourth trimester of 2021 (Fig. 1B). The average seroconversion rate in 2021 was 10.9% (95% CI 9.6 – 12.6) which is close to the rate of accumulated official reported COVID-19 cases in the city of Matinhos (11.5% in December 2021).

### IgG reactive to Nucleocapsid protein wane over time

Several studies indicate that IgG reactive to SARS-CoV-2 antigens wane over time, this reduction seems more rapidly for IgG reactive to the nucleocapsid antigen (Ortega *et al*. 2021). The first official COVID-19 case in the city of Matinhos date back to 30 March 2020 so it would be important to understand if the seroprevalence rate for the Nucleocapsid antigen could be underestimated by due to IgG wanning over the time. Longitudinal data from four Nucleocapsid seropositive subjects that participated in the study at least three times over >200 days are depicted in Fig. 2. The N protein reactive IgG signal waned over time for all investigated subjects (Fig. 2). In fact, subjects 1 and 2 were considered seroreverters > 200 after seroconversion as IgG levels dropped down below the assay cut off (Fig. 2). The identification of seroreverters during this study implies that the number of accumulated total COVID-19 cases in the population is likely to be underestimated in our serological survey (Fig. 1).

**Fig 2.**
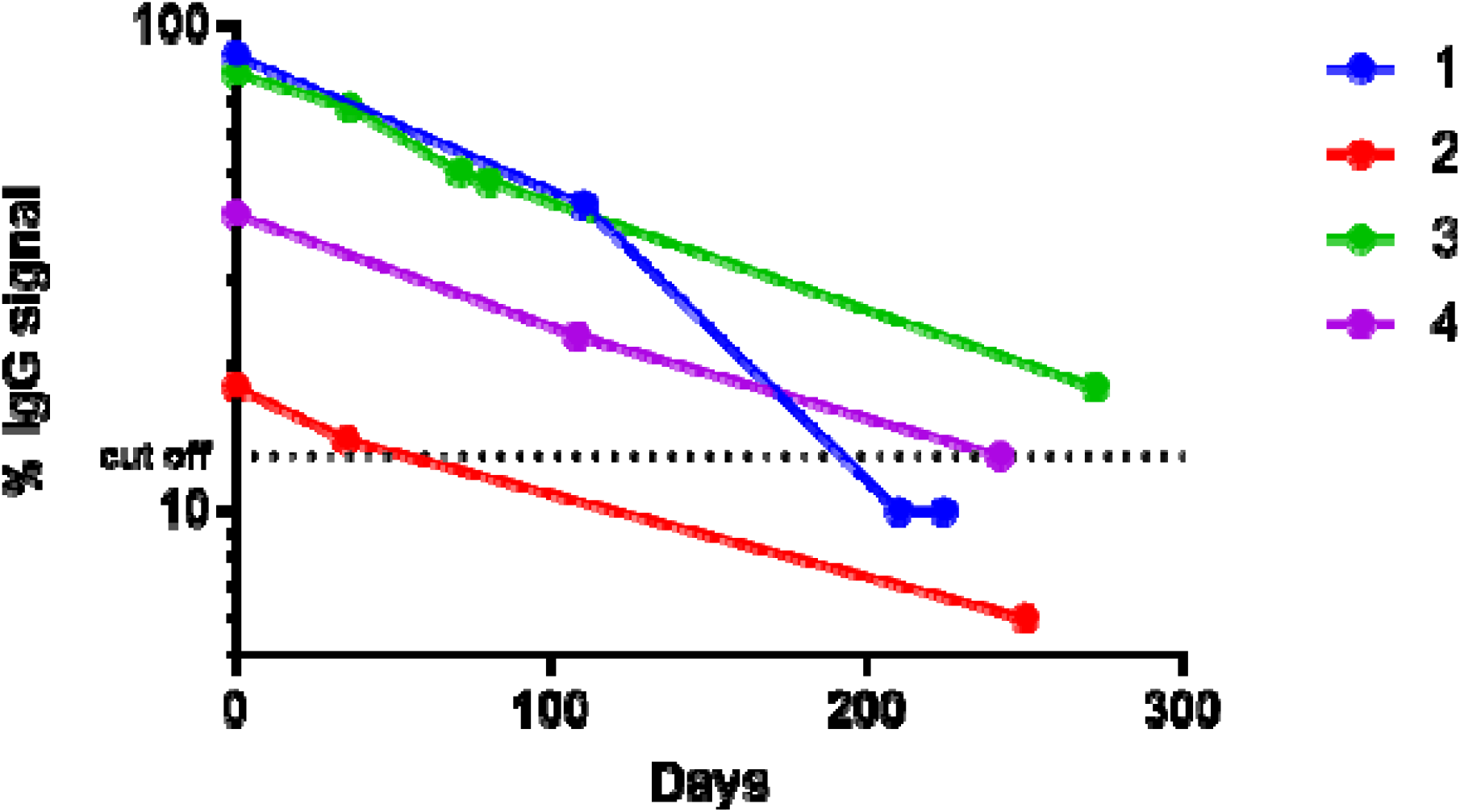
IgG reactive to Nucleocapsid antigen wane over time. Longitudinal analysis of IgG levels along the time in four subjects. IgG levels are reported as % of the assay calibrator, time zero was defined as the day of the detection of seroconversion. Subjects 1, 3 and 4 had at least two negative tests before detection of seroconversion. Subjects 2 and 3 had COVID-19 confirmed by qRT-PCR. Subjects 1 and 4 seroconverted few days after declaring contact with a household COVID-19 qRT-PCR confirmed case. Subject 1 was asymptomatic while the other subjects had presented mild symptoms.

### Serological response to immunization

The vaccine doses applied by October 2021 in Brazil were Pfizer-BioNTech 37.3%; AstraZeneca 36.6%; CoronaVac 24.5% and Janssen 1.6% https://www.gov.br/saude/pt-br/coronavirus/entregas-de-vacinas-covid-19. In order to detect the populational response to immunization, samples collected between July to November 2021 were analyzed for IgG reactive to full length Spike protein. By July 2021, seroconversion rates were 11% to Nucleocapsid and 30% to Spike antigen (Fig. 1 and 3). The difference in the Nucleocapsid to Spike seroconversion is likely to reflect the response to vaccination given that all vaccines would elicit the humoral response to the Spike antigen. The Spike seroconversion rates increased sharply reacting 76% in October 2021, a plateau is apparently being reached in November 2021 when 82% Spike seroconversion was detected (Fig. 3A).

**Figure 3.**
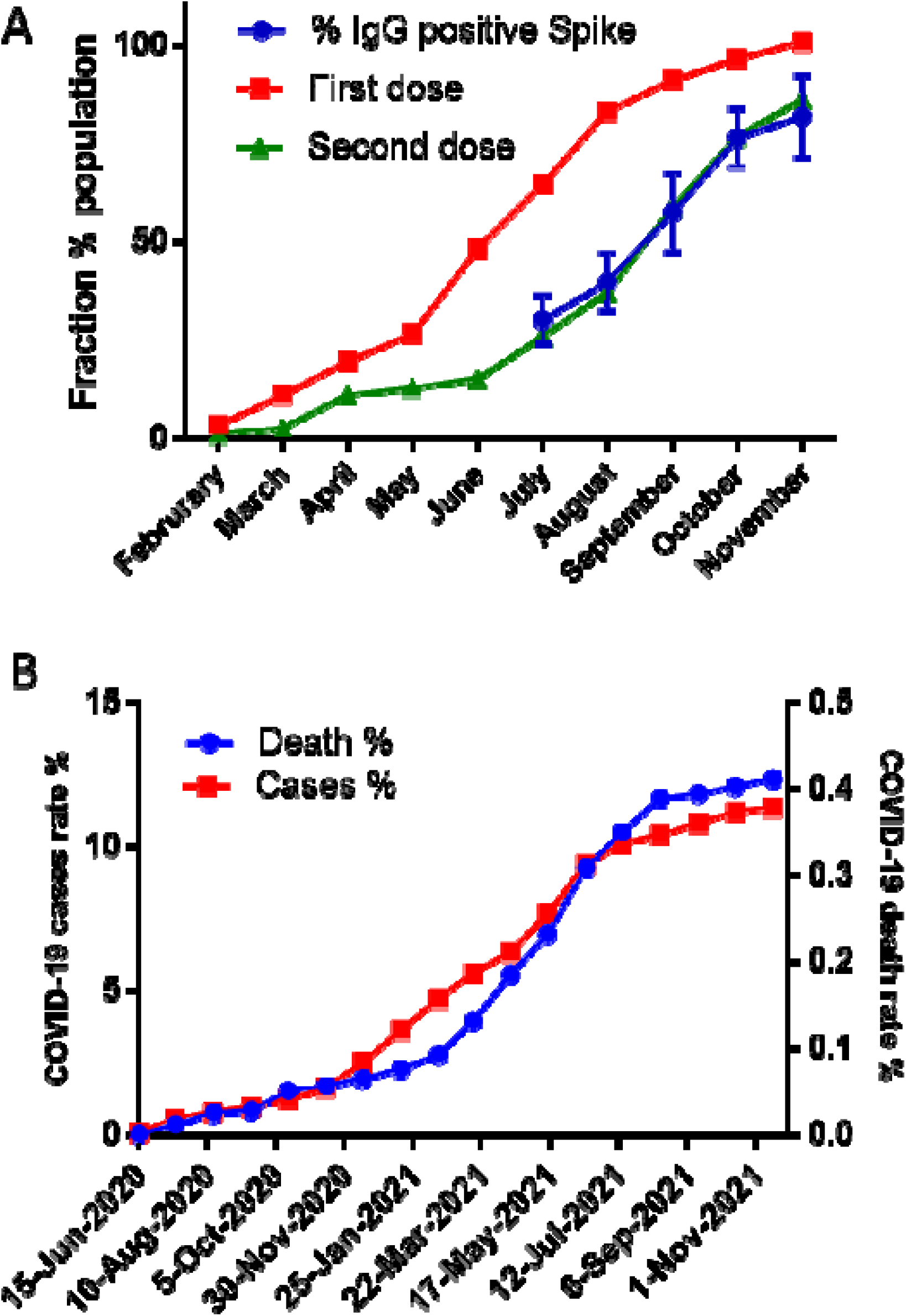
Evolution of vaccination and COVID-19 cases and deaths. A) The Fraction of the eligible (> 12 years old) vaccinated population with first dose (red), with the second dose (green) on the Paraná State. The rate of IgG seroconversion to Spike antigen detected in this study is shown in blue. Errors bars indicate the confidence intervals at 95% confidence level. B) Evolution of the rate of official COVID-19 cases (red) and death (blue) in the city of Matinhos.

The Spike seroconversion rates are well correlated with the official numbers of the second dose applied to the eligible population (> 12 years old) in the Paraná state (Fig. 3A). These data clearly indicate that the vaccines available in Brazil were successful the induction of the humoral response against the Spike antigen. The high level of vaccine coverage and seroconversion rates in the end of 2021 may explain the sharp decrease in COVID-19 accumulated deaths from September to December 2021 in the city of Matinhos (Fig. 3B).

It is worth mentioning that despite the full immunization with CoronaVac resulted in only 12% soroconversion for the Nucleocapsid antigen as mentioned earlier, up to 47% seroconversion was detected for the Spike antigen (considering samples collected > 10 days after the second dose and not declared to be previously diagnostic with COVID-19, N= 63). For those fully immunized with the Astrazeneca vaccine the seroconversion rate reached up to 75% >10 days after the second dose (N= 48). The number of samples analyzed after full immunization with Pfizer and Janssen were small (N < 10) and thus not considered for the rate of Spike seroconversion.

## Discussion

In this work we applied serological analysis to track SARS-CoV-2 seroconversion in response to infection and vaccination during the year of 2021 in the city of Matinhos, PR, Brazil. Seroconversion for the Nucleocapsid antigen was used to track previous infections after exclusion of CoronaVac vaccinated participants. On the other hand, Spike antigen seroconversion was used to track the humoral response to vaccination.

Seroconversion rates to Nucleocapsid antigen was not influenced by gender or age (Fig. 1A) which is in agreement with a previous study in Brazil (Hallal *et al*. 2020). The Nucleocapsid seroconversion rates were, in general, in agreement with official reported COVID-19 cases, especially during the second and third trimesters of 2021. More divergent numbers were observed in the 1^st^ and 4^th^ trimesters 2021 (Fig. 1B). We speculate that these discrepancies on the 1^st^ and 4^th^ trimester are result of case sub notification due to two different scenarios during the pandemic.

During the 1^st^ trimester of 2021, the worst phase of the pandemic was reached (Fig 3B), and availability of COVID-19 qRT-PCR tests were limited to severely symptomatic individuals. Investigation of case contacts were virtually nonexistent. The increase in molecular testing capacity during the year may explain why a better correlation between official COVID-19 cases and Nucleocapsid seroconversion rates were observed during the 2^nd^ and 3^rd^ trimesters (Fig. 1B). On the other hand, by the 4^th^ trimester of 2021, the vaccination cover was high and > 75% of the population had seroconverted to the Spike antigen (Fig. 3A). Vaccinated individuals are more likely to develop asymptomatic SARS-CoV-2 infections with lower viral load (Thompson *et al*. 2021; Franco-Paredes 2022) and hence not seek medical services or COVID-19 tests resulting in case sub notification. A plateau in the curve of official COVID-19 cases has been reached between the 3^rd^ and 4^th^ trimester 2021 (Fig. 3B). Our serological survey indicates a continuing growing trend in seroconversion in the same period (Fig. 1B) which suggests that SARS-CoV-2 continued to spread silentious in the population despite the high levels of immunization achieve by the vaccination campaign (Fig. 3A).

The number of official cases reported in the city of Matinhos by November 2021 was 4,052. If we consider the 16.7% seroconversion rate for Nucleocapsid antigen detected in November 2021, the number of total accumulated cases would be 5,962, suggesting a case sub notification in the range of 47% (1,910 cases). It is worth mentioning that technical constrains restrict the precision of the total cases that can be estimated with the serological survey. If one considers correction for assay sensitivity (95%) and specific (99.5%) the numbers of total cases is expected to increase by a factor of 4.5%, reaching up to 6,230 cases. Furthermore, it is important to note that seroreverters were detected during this study and thus the number of real COVID-19 infections should be even higher.

The official numbers for the infection fatality rate (IFR) by November 2021 in Matinhos was 3.6%. If we considered the case numbers obtained by our serological survey the IFR would be in the range of 2.4%. This number is closest to the IFR reported in Brazil and in the Paraná state of 2.7 and 2.6%, respectively (https://coronavirus.jhu.edu/data/mortality ; https://www.saude.pr.gov.br/Pagina/Coronavirus-COVID-19) but still higher than estimated by other serological surveys in Brazil were IFR in the range of 0.7 to 1% were estimated (Hallal *et al*. 2020; Marra and Quartin 2021). The differences in IFR numbers reported here and in other serological studies may be explained by the lack of correction for seroreversion rates. Furthermore, we speculate that local characteristics may result in higher IFR observed in the city of Matinhos, for instance, the human development index of Matinhos is below the national average and the city’s Hospital can deal only with low complexity cases.

The seroconversion rates for IgG reactive to Spike resembled the curve of the fraction the population that had taken the second vaccine dose (Fig. 3A). This data confirms that the vaccination campaign in Brazil was effective in activating humoral response at populational scale. The high vaccine coverage and high seroconversion rates are likely to be responsible for the reduction in official COVID-19 cases and deaths reported by the end of 2021 (Fig. 3B).

Our data unequivocally shows the success of the vaccination campaign to raise antibodies against the Spike antigen at the populational scale. The citizens and health authorities should be cautioned, however, because we did not investigate SARS-CoV-2 neutralizing antibodies in our analysis even though others have shown a correlation between the levels antibodies reactive to Spike and neutralizing activity (Ng *et al*. 2020). Furthermore, longitudinal studies are lacking so we cannot predict at this stage for how long the humoral response to vaccination is going to last.

Another point to consider is the emergence of SARS-CoV-2 variants such as B.1.1.529 (omicron) which has the potential to partially scape neutralizing antibodies raised after vaccination or after previous infections with the original SARS-CoV-2 (Planas *et al*. 2021). Since the first official case of B1.1.529 in the state Paraná in December 2021 https://www.aen.pr.gov.br/Noticia/Saude-confirma-primeiro-caso-da-variante-Omicron-no-Estado, the number of official cases COVID-19 are rapidly growing. Therefore, despite the high levels of seroconversion achieved by the vaccination campaign evidenced by this study (Fig. 3A) we strongly recommend that heath authorities continue to stimulate social distancing and use of face masks.

In conclusion, our SARS-CoV-2 seroconversion study in the city of Matinhos helped to understand the SARS-CoV-2 infection rates and the response to vaccination. It is likely that the numbers observed in the city of Matinhos may be similar to other similar cities in Brazil. Despite the investment in molecular testing facilities, much less attention has been given by heath authorities to deploy quantitative serological analysis. Serological analysis can help to track asymptomatic cases, case contacts, vaccine responses, and act as tool to distinguish COVID-19 from other diseases causing related symptoms such as dengue fever and influenza. We consider that the implementation of SARS-CoV-2 serological analysis as part of the public health services would provide key information to better manage the COVID-19 pandemic.

## Data Availability

All data produced in the present study are available upon reasonable request to the authors

## Acknowledgment

We acknowledgment the financial support of Alexander von Humboldt foundation, UFPR, CNPq, CAPES and Fundação Araucária. We are grateful to Prof. Karl Forchhammer (Tubingen University / Germany) and the Secretary of Heath of Matinhos for continuous support to this study. We acknowledge Profa Leda Castilhos, Laboratório de Engenharia de Cultivos Celulares (LECC) Coppe, UFRJ Rio de Janeiro, RJ, Brazil for providing the Spike protein used in this study.

